# First-in-human low-intensity focused ultrasound targeting striatal circuits in schizophrenia: feasibility, safety, and effects on hallucinations and striatal-temporal functional connectivity

**DOI:** 10.64898/2026.01.10.26343837

**Authors:** Karuna Subramaniam, Grace Attalla, Miriam Mathew, John L. Alvarez, Ehsan Dadgar-Kiani, Rajiv Mahadevan, Srikantan Nagarajan, Keith R Murphy

## Abstract

**Background:** Auditory hallucinations are among the most disabling symptoms in individuals with schizophrenia (SZ) and are linked to aberrant signaling within deep-striatal circuits, such as the nucleus accumbens (NAc) and caudate head (CH). However, causal tests of striatal involvement have been limited by the inaccessibility of these structures using noninvasive neuromodulatory techniques. Low-intensity focused ultrasound (LIFU) provides millimeter-scale precision capable of modulating deep-brain circuits, but its feasibility and impact on hallucinations in SZ remain unknown.

**Methods:** SZ participated in a within-subject cross-over feasibility trial including two active LIFU sessions (NAc, CH) and one sham control (unfocused sonication), spaced one-week apart. Resting-state fMRI and hallucination symptoms were acquired at baseline and immediately post-sonication.

**Results:** LIFU was delivered safely and well-tolerated in all patients. Acoustic simulations show consistent engagement of both striatal targets across subjects. Clinically, SZ demonstrated significant reductions in hallucination severity following active LIFU to NAc and CH, relative to baseline. Mechanistically, SZ exhibited abnormally high striatal–superior temporal cortex (STC) connectivity at baseline. Immediately after sonication, active LIFU to NAc and CH produced robust reductions in striatal–STC coupling in SZ.

**Conclusions:** This first-in-human study demonstrates that deep striatal LIFU is safe, feasible, and produces functional-connectivity changes accompanied by hallucination severity reductions in SZ. The convergence of symptom improvement with reduced striatal–STC coupling provides mechanistic proof-of-concept evidence that this circuit provides a promising biomarker and therapeutic LIFU target in psychosis and motivates larger sham-controlled trials to test the causal role of striatal circuitry in hallucination generation in SZ.

## INTRODUCTION

Schizophrenia is a serious neuropsychiatric disorder affecting approximately 1% of the population worldwide. Auditory hallucinations are the most common, persistent, and disabling psychotic symptom in individuals with schizophrenia (SZ), affecting over 70% of patients (1). These hallucinations typically involve hearing critical or threatening voices in the absence of external auditory input, and are associated with profound functional impairment and increased mortality(2). Dysfunctional neural signaling in key striatal subregions, the nucleus accumbens (NAc) and caudate, has been strongly implicated in their pathogenesis, with studies showing hyperactivity, increased neural connectivity and excessive dopamine (DA) release in these regions(3–24). Current antipsychotic medications exert their effects by acutely blocking DA D_2_ receptors and gradually reducing striatal DA release over several weeks (14, 25). However, these medications remain inadequate with symptoms persisting in up to 40% of patients (26). Even clozapine, which is considered the most effective antipsychotic treatment for refractory SZ, yields only partial response in 40–70% of patients and causes substantial side effects due to diffuse off-target circuit engagement (26, 27). These limitations underscore the urgent need for mechanistic approaches to test how striatal dysfunction causally contributes to auditory hallucinations.

Auditory phantom percepts such as hallucinations are hypothesized to arise from aberrant functional connectivity between the NAc/CH and superior temporal gyrus (STC), which disrupts information-gating mechanisms. This dysregulated gating of information is thought to allow internally-generated thoughts and inner speech to be misperceived as external, giving rise to the experience of auditory hallucinations (5, 6, 28–31). The STC, including Heschl’s gyrus (HG) and planum temporale (PT), are activated in healthy participants during perception of external auditory stimuli such as speech (32–35). In contrast, SZ show volumetric deficits in the STC and exhibit abnormally increased activation during auditory hallucinations, even in the absence of external auditory input(6, 36–43). Furthermore, SZ who experience hallucinations show abnormally elevated functional connectivity involving the striatum (NAc, CH) and STC, compared to non-hallucinating SZ and healthy participants(5, 6, 14, 30). This hyperconnectivity indicates that striatal-STC coupling is a central circuit-level mechanism underlying the pathophysiology of hallucinations in SZ. While invasive deep brain stimulation (DBS) of the NAc or caudate can reduce psychotic symptoms (7–11, 13-(14), its clinical use is limited by surgical risk and sustained physical and psychiatric adverse effects in SZ(7–11, 13). In contrast, low-intensity focused ultrasound (LIFU) offers a safe non-invasive alternative capable of modulating deep-brain circuits with millimeter-level spatial and sub-second temporal precision (44–52). LIFU’s favorable safety profile and capacity for real-time physiological monitoring make it uniquely suited to probe the causal role of striatal circuits in SZ (44–51).

Here, we present the first-in-human feasibility data applying LIFU to the NAc and CH in SZ. Using individualized MRI-based targeting and neuronavigation, we establish proof-of-concept findings for future sham-controlled studies testing whether modulation of striatal circuitry can causally influence auditory hallucinations in SZ. These data demonstrate the safety, tolerability and feasibility of striatal LIFU in this population, and explore early neural and behavioral signatures of striatal target engagement. This proof-of-concept study had three primary objectives: (i) To establish the safety and feasibility of striatal target engagement in SZ. We hypothesized that all sessions would be well-tolerated, and that subject-specific acoustic simulations would demonstrate robust spatial-peak pulse-average intensities (ISPPA) precisely within the intended NAc and CH targets. (ii) To evaluate effects of LIFU on auditory hallucinations in SZ. We hypothesized that stimulation of NAc and CH would induce short-term reductions in hallucination severity immediately following sonication, and (iii) To identify neural mechanisms by which LIFU may influence hallucinations in SZ. We hypothesized that LIFU would produce measurable modulation of striatal–temporal circuitry, consistent with mechanistic models of hallucination generation.

## METHODS

### Participants

This study was conducted under an NIMH-funded R01 (R01MH122897) to Karuna Subramaniam and designated non-significant risk (NSR) by the UCSF Institutional Review Board (IRB #24-42726). SZ were recruited via ClinicalTrials.gov (NCT04807530) and provided informed consent for this protocol approved by Attune Neurosciences and the IRB at UCSF prior to study enrollment. All procedures were conducted at UCSF. SZ participants (n=3) then completed this study and all clinical assessments at UCSF (**Table 1** summarizes demographic and clinical characteristics).

**Table 1.**
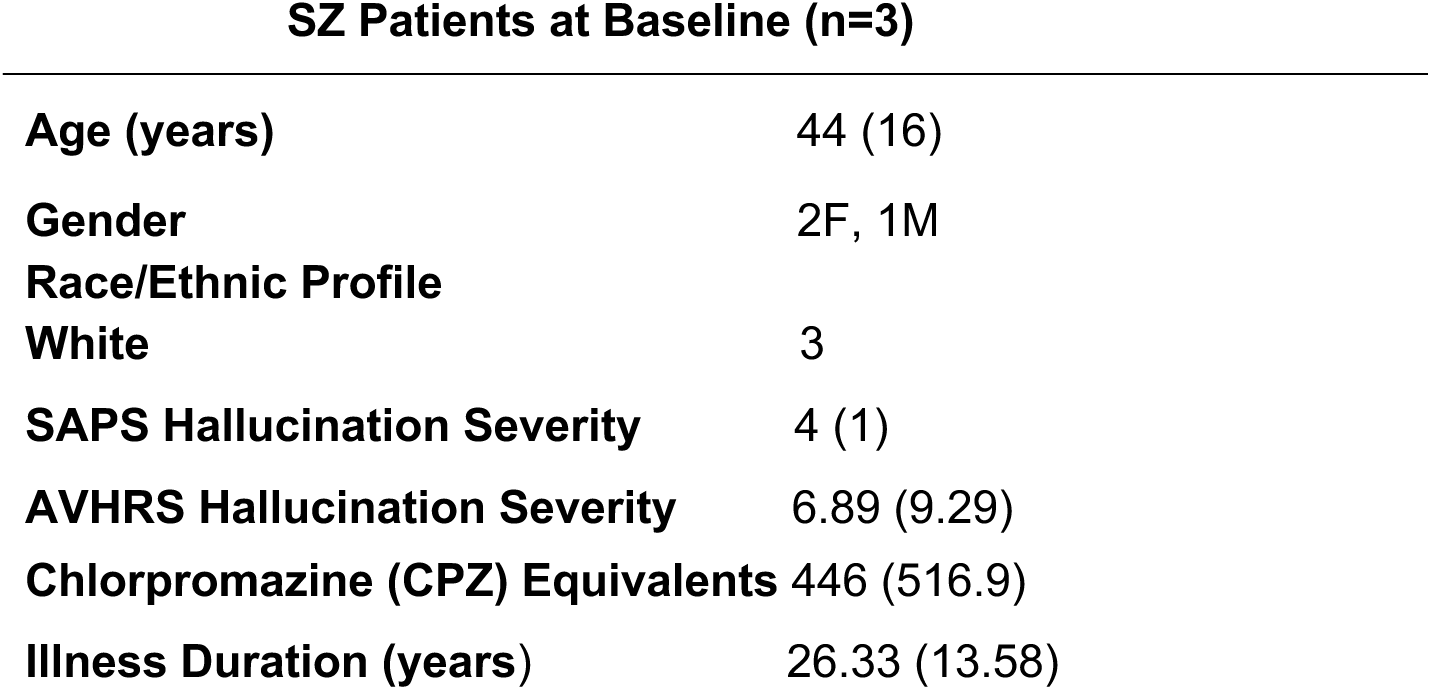
Demographic and Clinical Profiles (mean, SD) of Schizophrenia Participants (SZ) SZ Patients at Baseline (n=3)

| SZ Patients at Baseline (n=3) |  |
| --- | --- |
| Age (years) | 44 (16) |
| Gender | 2F, 1M |
| Race/Ethnic Profile |  |
| White | 3 |
| SAPS Hallucination Severity | 4 (1) |
| AVHRS Hallucination Severity | 6.89 (9.29) |
| Chlorpromazine (CPZ) Equivalents | 446 (516.9) |
| Illness Duration (years) | 26.33 (13.58) |

### Study Design

This first-in-human (FIH), sham-controlled randomized within-subject cross-over Phase 1 feasibility trial included five in-person visits. Visit 1-2 included baseline visits (diagnostic evaluation, MRI/fMRI acquisition, neurological exam and clinical assessments). Visits 3–5 each included one of the three LIFU sessions given in randomized order, spaced one week apart: (i) Active LIFU targeting NAc, (ii) Active LIFU targeting CH, and (iii) Sham control using unfocused sonication (UF). Daily follow-up clinical assessments were conducted for one week following each LIFU session to monitor safety and symptom changes.

### Inclusion Criteria

Inclusion criteria were: 1) Participants with DSM-V schizophrenia diagnosis (SCID) experiencing auditory hallucinations; 2) age 18-64; 3) stable outpatient status ≥12 weeks on unchanged medications ≥4 weeks; (4) no neurological or major medical illness affecting neural function; (5) no current substance abuse; (6) no MRI contraindications; and (7) negative pregnancy test for females.

### Clinical Measures

Hallucination severity was assayed using the Visual Analog Scale (VAS) and Auditory Vocal Hallucination Rating Scale (AVHRS) (53–55). The VAS (0 = “Not at all severe” to 100 = “Very severe”) served as the primary high-resolution measure providing sensitive daily hallucination symptom tracking, and capturing acute hallucination changes before, during and after each LIFU session. AVHRS served as a validated index of baseline hallucination severity and cumulative symptom change over the one-week post-LIFU period, complementing the VAS as a sensitive measure of acute hallucination symptom changes. VAS ratings were collected daily for one week pre-LIFU to confirm stability prior to each LIFU condition, as well as immediately before, during, and after each LIFU session, and daily for one week post-LIFU. AVHRS was administered at baseline and at one week post-sonication. Both measures demonstrate high reliability (ICCs ≥ 0.80)(56, 57),(58–60), and are well-established for detecting short-term and sustained hallucination symptom changes in psychosis.

### Imaging Acquisition and Functional Connectivity Analyses

MRI data were acquired on a 3T Siemens Prisma MRI scanner with 64- and 20-channel head-neck coils. High-resolution anatomical scans were obtained using T1-weighted MPRAGE (TR=2300 msec, TE=3.03 msec, 208 slices, .94mm slice thickness) and rs-fMRI (eyes open) using multi echo-planar imaging (TR=607ms, TE=32ms, 65 slices, 2.5mm slice thickness). Preprocessing and connectivity analyses used SPM12 and CONN v22. Seed-to-voxel connectivity analyses (Fisher Z-transformed correlations) were performed between striatal seeds (NAc, CH) and STC regions implicated in hallucination-related striatal–temporal hyperconnectivity (5, 6, 14, 30).

### LIFU Targeting and Stimulation

Individualized MRI-based neuronavigation was performed using Attune software which integrated each patient’s T1-weighted (MPRAGE) and pseudo-CT (PETRA) images to estimate skull-specific acoustic properties for precise LIFU targeting. Skull density and speed of sound values were assigned using an established mapping from Siemens PETRA ultra-short echo time MRI scans to Hounsfield units based on Miscouridou et al (61). Simulation output was calibrated to the device in 3D steerable space as described previously (62). Simulations were performed at the resolution of the PETRA scans (0.7 mm), with a homogenous bone attenuation assignment of 10.3 dB/cm based on our previous study with 8 temporal window skull fragments (62). Thermal modeling was performed with k-wave using Pennes bioheat equation, assuming absorption represented 16% of attenuative losses (63). Thermal simulations assumed a static focus despite lateral shifts. NAc and CH coordinates (e.g., NAc: −8, 9, −8; CH: −10, 18, 2 in MNI space) were transformed into native anatomy. LIFU was delivered using the Attune ATTN201 system, operating at 500 kHz with bilateral 64-element transducer arrays. Session sonications were composed of 20 Hz pulse repetition frequency, 6% duty cycle (3 ms pulses, 50 ms interval), applied in 40 s on / 20 s off cycles for 30 minutes per hemisphere (60 minutes total) (**Supplemental Fig. 1**). Element driving voltage and resulting source pressures were fixed for all stimulations irrespective of target or anatomical differences to best equate dosing and peripheral confounds across treatments. Sham sessions used unfocused sonication which randomizes delays for each target to produce negligible focal pressure, while delivering the same total ultrasound energy (**Supplemental Methods**). Device placement accuracy was ensured by use of an augmented reality projection of the device position in addition to mechanical registration tools including a nose bridge fit tool and custom headband used across sessions.

### Safety and Monitoring

All expected acoustic outputs were within the International Transcranial Ultrasonic Stimulation Safety and Standards Consortium (ITRUSST) guidelines (64–66). All 95^th^ percentile error-tolerance estimates for brain mechanical index (MI) were below 1.9. No adverse events of any kind were associated with the stimulation session. Simulated maximum temperature rise in all tissue types was ≤ 0.6 °C, well below the ITRUSST recommended limit (Table 2). All sessions were well-tolerated; no participant reported more than mild, transient auditory sensations (e.g. brief auditory clicks) as expected and described in previous works.

**Table 2.** Stimulus safety estimates.

| Subject | Target | Target Peak Pressure (MPa) | Target ISPPa (W/cm²) | Target Peak Pressure 5th %ile (MPa) | Target Peak Pressure 95th %ile (MPa) | Target Temp Rise (°C) | MI Brain | MI Brain 5th %ile | MI Brain 95th %ile | MI Head | MI Skull | MI Soft Tissue | TR Brain (°C) | TR Skull (°C) | TR Soft Tissue (°C) |
| --- | --- | --- | --- | --- | --- | --- | --- | --- | --- | --- | --- | --- | --- | --- | --- |
| SZ01 | UnfLeft | 0.04 | 0.04 | 0.02 | 0.05 | 0.0 | 0.23 | 0.13 | 0.31 | 0.74 | 0.74 | 0.65 | 0.11 | 0.22 | 0.19 |
| SZ01 | UnfRight | 0.07 | 0.17 | 0.04 | 0.1 | 0.01 | 0.34 | 0.19 | 0.46 | 1.22 | 0.94 | 1.22 | 0.21 | 0.42 | 0.37 |
| SZ01 | NAcRight | 0.43 | 5.6 | 0.24 | 0.57 | 0.08 | 0.63 | 0.35 | 0.84 | 1.28 | 1.28 | 1.27 | 0.36 | 0.61 | 0.5 |
| SZ01 | NAcLeft | 0.43 | 5.58 | 0.24 | 0.57 | 0.09 | 0.62 | 0.35 | 0.82 | 1.29 | 1.29 | 1.29 | 0.42 | 0.71 | 0.61 |
| SZ01 | CaudHeadLeft | 0.56 | 9.8 | 0.32 | 0.75 | 0.15 | 0.8 | 0.45 | 1.06 | 1.17 | 1.17 | 1.15 | 0.41 | 0.73 | 0.6 |
| SZ01 | CaudHeadRight | 0.56 | 9.58 | 0.31 | 0.74 | 0.14 | 0.79 | 0.44 | 1.05 | 1.2 | 1.19 | 1.2 | 0.32 | 0.65 | 0.55 |
| SZ02 | NAcRight | 0.65 | 13.14 | 0.37 | 0.87 | 0.19 | 0.93 | 0.52 | 1.23 | 1.45 | 1.45 | 1.3 | 0.41 | 0.69 | 0.56 |
| SZ02 | NAcLeft | 0.65 | 13.11 | 0.36 | 0.87 | 0.19 | 0.93 | 0.52 | 1.23 | 1.53 | 1.53 | 1.44 | 0.43 | 0.7 | 0.59 |
| SZ02 | CaudHeadRight | 0.61 | 11.45 | 0.34 | 0.81 | 0.16 | 0.86 | 0.48 | 1.15 | 1.4 | 1.4 | 1.23 | 0.42 | 0.67 | 0.56 |
| SZ02 | CaudHeadLeft | 0.62 | 11.74 | 0.35 | 0.82 | 0.17 | 0.87 | 0.49 | 1.16 | 1.6 | 1.6 | 1.19 | 0.4 | 0.72 | 0.56 |
| SZ02 | UnfRight | 0.09 | 0.27 | 0.05 | 0.12 | 0.01 | 0.58 | 0.33 | 0.78 | 1.26 | 1.26 | 1.23 | 0.28 | 0.44 | 0.4 |
| SZ02 | UnfLeft | 0.04 | 0.05 | 0.02 | 0.05 | 0.01 | 0.73 | 0.41 | 0.97 | 1.41 | 1.41 | 1.0 | 0.3 | 0.55 | 0.41 |
| SZ03 | CaudHeadLeft | 0.69 | 14.62 | 0.39 | 0.92 | 0.2 | 0.97 | 0.55 | 1.29 | 1.27 | 1.27 | 1.22 | 0.38 | 0.61 | 0.52 |
| SZ03 | CaudHeadRight | 0.6 | 11.29 | 0.34 | 0.8 | 0.17 | 0.86 | 0.48 | 1.14 | 1.55 | 1.55 | 1.29 | 0.34 | 0.71 | 0.58 |
| SZ03 | UnfRight | 0.06 | 0.11 | 0.03 | 0.08 | 0.01 | 0.58 | 0.32 | 0.77 | 1.04 | 0.91 | 1.04 | 0.16 | 0.42 | 0.36 |
| SZ03 | UnfLeft | 0.03 | 0.03 | 0.02 | 0.04 | 0.01 | 0.78 | 0.44 | 1.04 | 0.97 | 0.97 | 0.88 | 0.18 | 0.28 | 0.25 |
| SZ03 | NAcLeft | 0.83 | 21.47 | 0.47 | 1.11 | 0.3 | 1.19 | 0.66 | 1.58 | 1.2 | 1.2 | 1.2 | 0.32 | 0.66 | 0.58 |
| SZ03 | NAcRight | 0.73 | 16.35 | 0.41 | 0.97 | 0.24 | 1.08 | 0.6 | 1.43 | 1.26 | 1.2 | 1.26 | 0.32 | 0.7 | 0.63 |

### Statistical Analysis

Feasibility and tolerability outcomes were summarized descriptively across sessions. Given the small sample (n=3), statistical analyses were exploratory and focused on estimating effect direction and magnitude rather than formal inference. Repeated-measures ANOVAs assessed main effects of sonication condition, and paired t-tests evaluated within-subject changes in hallucination severity and frequency (VAS) from baseline. Changes in striatal–temporal connectivity (Fisher Z–transformed correlations; range = –1 to +1) were visualized to illustrate the magnitude and directionality of modulation following active sonication versus baseline.

## RESULTS

### LIFU Target Engagement in SZ

Using calibrated simulations validated by through-skull measurements, we observed focal overlap at both individualized striatal targets (**Fig. 1a-b**) (67) with target estimated spatial-peak pulse-average intensity (ISPPA) of 7.72-22.86 W/cm² at the NAc (MNI −8, 9, −8) and 12.47-15.77 W/cm² at the CH (MNI −10, 18, 2) (**Fig. 1c**). The UF sham condition produced no focal pressure hotspots, validating it as a credible control and had a ISPPA range of 0.03 - 0.28 W/cm² (**Fig. 1c**). All sessions were well-tolerated. Predicted acoustic indices remained under the ITRUSST safety thresholds (Table 2; MI<1.9; ΔT<2°C at skull; <1°C in situ) (64–66). Together, these findings establish the safety, tolerability, and feasibility of targeting deep striatal structures in SZ using neuronavigated LIFU.

**Figure 1.**
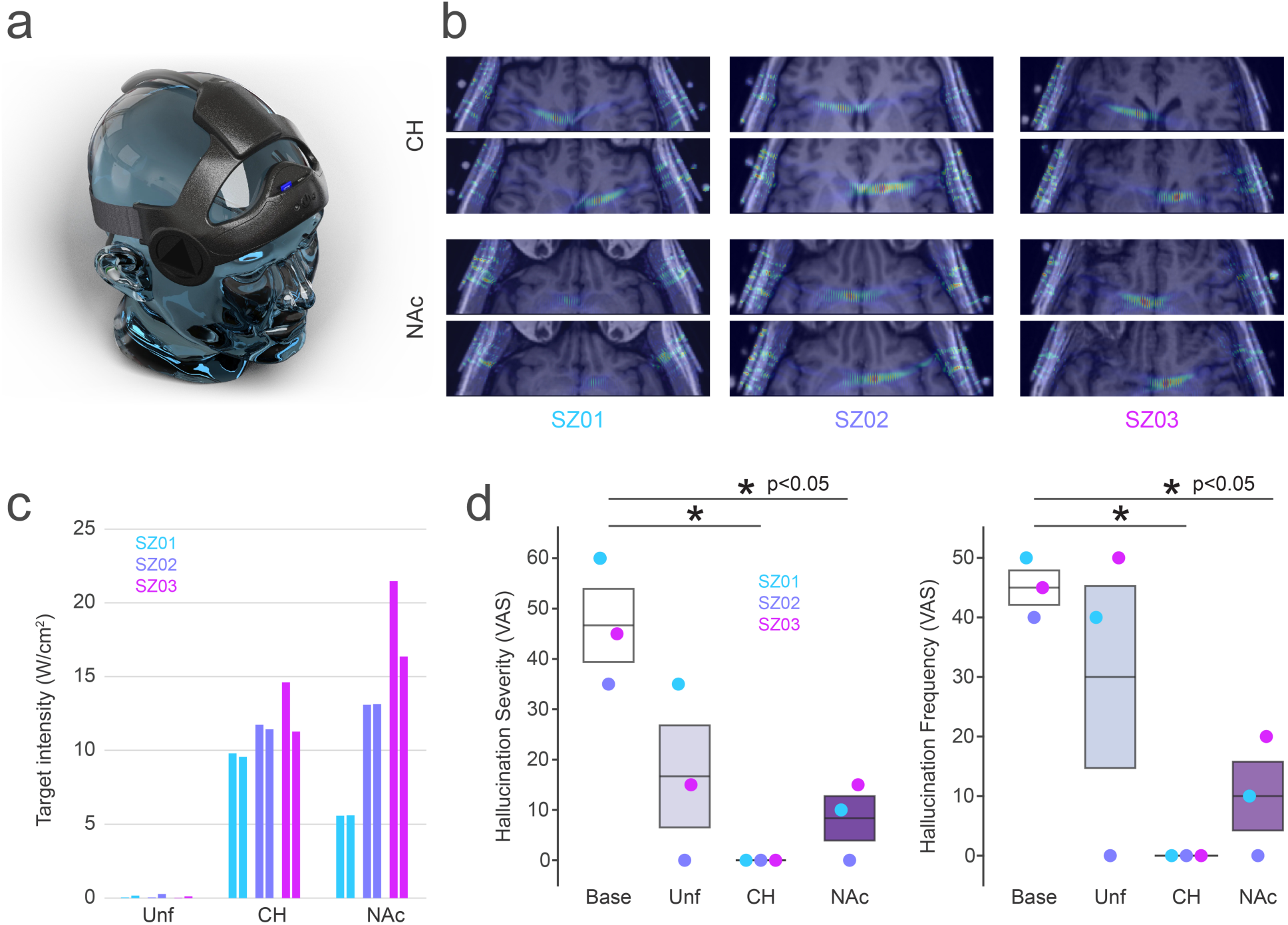
Acoustic simulations demonstrate precise striatal target engagement and reveal acute reductions in hallucination severity and frequency following LIFU. **a-b.** Validated through-skull simulations produced robust focal overlap at both individualized striatal targets. **c.** Acoustic intensity maps showed a target situ ISPPA of 7.72 - 22.86 W/cm² at NAc (MNI −8, 9, −8) and 12.47 - 15.77 W/cm² at CH (MNI −10, 18, 2), and lower intensities (∼6-7 W/cm²) in adjacent off-target regions. The UF sham condition produced no focal brain pressure hotspots, validating it as a credible control and had a ISPPA range of 0.03 - 0.28 W/cm². **d.** Box plot lines depict mean hallucination severity and frequency ratings (VAS) at baseline and immediately after sonication across conditions. Box plots bars show group ± standard error of the mean; colored dots denote individual patients (SZ 01–03). Both active LIFU conditions (CH, NAc) produced significant reductions in hallucination severity and frequency relative to baseline (*p < .05). SZ 02 reported no hallucinations immediately prior to the UF control session, precluding a pre-post comparison for that condition. Aside from this case, pre-sonication ratings were consistent with each patient’s baseline values. Active LIFU yielded >50% reductions in hallucination severity and frequency in all patients, relative to baseline.

### LIFU-Induced Reductions in Hallucination Severity and Frequency

Hallucination data were collected daily for one week prior to each LIFU session to establish stable baseline levels in all three patients. VAS hallucination ratings were collected immediately before and after each LIFU sonication session. A repeated-measures ANOVA demonstrated a significant main effect of sonication condition on hallucination severity (F=17.2, p=.002) and frequency (F=8.5, p=.01). Follow-up paired comparisons showed robust pre-post reductions in hallucination severity following active LIFU to NAc (t(2)=6.38, p=.02) and CH (t(2)=6.42, p=.02) and in hallucination frequency after LIFU to NAc (t=7.0, p=.02) and CH (t=15.6, p=.004) (**Fig. 1d**).

Across all participants, active LIFU to NAc and CH produced large, clinically meaningful reductions, with every patient showing >50% decrease in both hallucination severity and frequency relative to baseline (**Fig. 1d**). Due to the absence of hallucinations prior to the UF condition in one patient (SZ 02), statistical comparisons involving the UF condition were limited. Visual inspection nonetheless indicated lower average post-sonication hallucination ratings following active LIFU (NAc and CH) than UF. Given the very small sample, effect sizes are provided for descriptive context only and should be interpreted cautiously: Cohen’s d = 0.58 (NAc vs. UF) and d = 0.95 (CH vs. UF) for severity.

Hallucination symptoms returned to baseline by the end of each washout period prior to subsequent active LIFU sessions, supporting adequate washout for evaluating active stimulation effects. Blinding integrity was strong: participants were informed only that “the study examines the effects of brain stimulation to different regions linked to hallucinations,” and none were aware of the sham condition. During debriefing, no patient correctly identified the UF session, and all reported similar sensory experiences, stating the sessions “felt the same,” confirming successful patient blinding.

### LIFU-Induced Modulation of Striatal–Temporal Connectivity

Resting-state fMRI was obtained at baseline and immediately after each active LIFU session to assess LIFU-induced neural effects. One patient was unavailable for post-LIFU imaging due to scheduling constraints. At baseline, all patients showed abnormally elevated functional connectivity between striatal seeds (NAc, CH) and the STC, particularly within Heschl’s gyrus (HG) and the planum temporale (PT) (**Fig. 2**). These results are consistent with prior reports linking striatal–STC hyperconnectivity to auditory hallucinations in SZ (5, 6, 14, 30).

**Figure 2.**
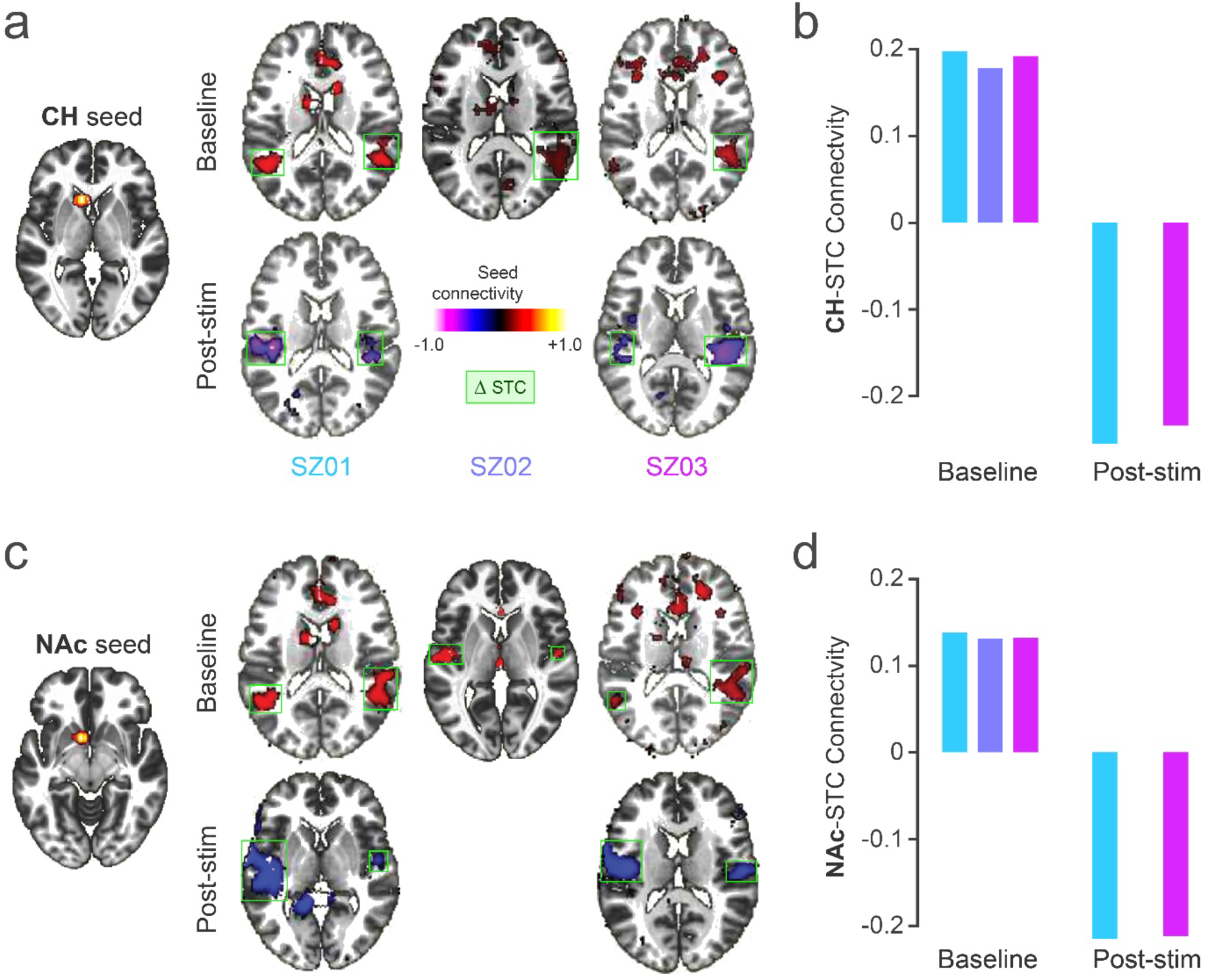
Modulation of Aberrant Striatal–Temporal Connectivity following Striatal LIFU in SZ. Baseline seed-to-voxel connectivity maps show abnormally increased connectivity between **(a-b)** CH and **(c-d)** NAc seeds with STC (red clusters) in SZ (n=3), particularly including auditory temporal sites in HG and PT (peak coordinates: −40, −26, 14 and 56, −30, 14). **b, d.** Baseline Fisher Z values demonstrate consistent hyperconnectivity across all patients. Following neuronavigated LIFU to **(a-b)** CH and **(c-d)** NAc, SZ exhibited robust decreases in striatal-STC connectivity (blue clusters), particularly within HG and PT (peak coordinates: −48, −22, 12 and 50, −18, 12). **b, d.** Post-LIFU Fisher Z values demonstrate reduced striatal-STC coupling relative to baseline, indicating modulation of this aberrant circuitry implicated in hallucination generation in SZ.

Following active LIFU, both NAc- and CH-targeted sonications produced marked reductions in striatal–STC connectivity (**Fig. 2**), relative to baseline. Connectivity reductions following LIFU to NAc and to CH yielded medium effect sizes (Cohen’s *d* = 0.43–0.52), consistent with meaningful modulation of striatal–temporal coupling in this first-in-human sample. These connectivity reductions were observed within auditory temporal regions, particularly within HG and PT sites, which are strongly implicated in the pathogenesis of hallucinations in SZ (6, 36–43). These initial findings demonstrate the feasibility of using LIFU to modulate aberrant striatal–temporal coupling—a circuit-level mechanism implicated in hallucination generation in SZ.

### Summary

These first-in-human data demonstrate that active striatal LIFU produced rapid, robust and clinically meaningful reductions in hallucination severity and frequency in every patient, with clear pre–post effects at both striatal targets and appropriate washout between sessions. These findings provide proof-of-concept evidence that LIFU can achieve targeted, circuit-specific modulation of psychotic symptoms. We further demonstrate that LIFU can safely and precisely engage deep striatal targets (NAc, CH) in SZ and acutely modulate aberrant striatal–temporal connectivity, a circuit strongly implicated in auditory hallucinations. Together, these results identify the striatal–STC pathway as a promising biomarker and a mechanistically-relevant target for noninvasive neuromodulation in psychosis. Overall, these initial findings support the feasibility, tolerability, neural specificity and symptom relevance of deep striatal LIFU in SZ and provide a compelling rationale for larger sham-controlled studies to test the causal role of striatal–temporal circuitry in the generation of auditory hallucinations.

## DISCUSSION

This first-in-human feasibility study provides initial evidence that LIFU can safely, precisely, and reversibly modulate deep striatal circuits implicated in the generation of auditory hallucinations in SZ. Across three SZ patients with hallucinations, neuronavigated LIFU to the NAc and CH produced: (i) reliable target engagement confirmed by robust ISPPA acoustic simulations localized precisely within the intended NAc and CH targets; (ii) acute, clinically meaningful reductions in hallucination severity and frequency; and (iii) modulation of aberrant striatal–STC functional connectivity. Together, these findings provide mechanistic proof-of-concept evidence that aberrant striatal–STC coupling is not only a biomarker of hallucinations but also a *modifiable* neural substrate. More broadly, they suggest that deep-brain LIFU may represent a promising novel circuit-directed therapeutic approach for psychosis.

### Safety and Feasibility of Deep Striatal LIFU

The present work demonstrates that bilateral striatal LIFU can be delivered safely and with high tolerability in SZ—a population in whom invasive procedures pose heightened risk (7–11, 13-(14). All acoustic parameters conformed to FDA and ITRUSST guidelines (64–66), with mechanical index and temperature rise well below safety thresholds. No participant experienced any adverse events beyond mild, transient sensations consistent with prior tFUS neuromodulation studies using comparable sonication intensities in which no adverse event was observed(47, 48, 68). Importantly, the sham condition (unfocused sonication) produced no focal brain energy deposition, validating its use as a credible control in future trials.

These results extend prior LIFU studies targeting cortical and subcortical targets across healthy and neurological populations (44–52, 69–71) by demonstrating safe, precise delivery to deeper, clinically relevant striatal foci in SZ, a group historically underrepresented in neuromodulation research. To our knowledge, this is the first proof-of-concept evidence that striatal LIFU is safe, well-tolerated, and feasible for clinical translation in SZ.

### Symptom Reductions Following Striatal LIFU

Active LIFU to both striatal targets yielded rapid and substantial reductions in hallucination severity and frequency, with effects emerging during stimulation and persisting during the immediate post-sonication period. All participants showed >50% reduction in hallucination severity following both NAc and CH stimulation, with effect sizes in the medium-to-large range and clear separation from baseline levels. Although direct statistical comparisons with the sham condition were limited by the lack of hallucination symptoms prior to the unfocused sonication in one participant, visual inspection revealed a pattern of lower average post-sonication hallucination ratings following active LIFU compared to sham.

These results converge with reports that DBS to the NAc or caudate can ameliorate psychotic symptoms (7–11, 13–14), but importantly, LIFU achieves these effects noninvasively and with minimal risks. The rapid time course of symptom change further supports the hypothesis that LIFU modulates neural circuit function directly, rather than through delayed neurochemical effects as seen with pharmacologic agents (14, 25). If replicated in larger samples, LIFU could offer a mechanistically targeted intervention for hallucinations with advantages over both antipsychotic medication that cause substantial side effects due to diffuse off-target circuit engagement (26, 27) and multiple sessions of surface-based brain neuromodulation TMS approaches that cannot directly access deep striatal circuitry.

### Striatal Neuromodulation and Psychosis: Implications for Causal Circuit Mechanisms

The striatum—particularly the NAc and CH—has long been implicated in schizophrenia via converging evidence from dopamine imaging, postmortem studies, electrophysiology, and connectivity analyses (3–24). Hyperdopaminergia, elevated intrinsic activity, aberrant excitatory input, and increased coupling with auditory temporal areas have all been associated with hallucination severity (5, 6, 14, 30). Yet despite this extensive correlational literature, causal evidence linking striatal dysfunction to hallucination generation remains scarce, due in part to the inaccessibility of deep subcortical structures using traditional noninvasive neuromodulation.

Our findings demonstrate that LIFU can transiently reduce the abnormal striatal–STC hyperconnectivity in hallucinating SZ and that these connectivity changes emerge in concert with acute hallucination symptom improvement. While based on a small first-in-human sample, this pattern aligns with mechanistic models positing that excessive striatal drive may bias STC circuits toward misperceiving the source of self-generated inner experiences as external auditory input (e.g. hearing voices when there are none), resulting in hallucinations (28, 72–76). By attenuating excessive striatal–STC coupling (5, 6, 14, 30), LIFU may help restore the balance between internally-generated auditory predictions and externally-driven auditory signals, thereby reducing hallucination symptoms.

These observations align with deep brain stimulation (DBS) reports describing symptom reductions when stimulating NAc or caudate targets in treatment-resistant SZ (7–11, 13–14). Unlike DBS, however, LIFU provides a reversible, low-risk method for testing causal circuit mechanisms in psychosis with millimeter precision (44–52). The present study therefore advances the emerging field of deep ultrasonic neuromodulation into a critical neuropsychiatric domain where mechanistic interventions has historically been limited.

### Limitations and Future Directions

As a feasibility study, the sample was intentionally small and exploratory, and therefore not designed to establish definitive clinical efficacy. The sample size (n=3) appropriately constrains generalizability and the strength of statistical inference, yet provides critical preliminary insights to guide future, adequately powered trials. Resting-state fMRI was obtained post-LIFU in two SZ patients, both of whom showed convergent reductions in striatal–STC connectivity across targets. However, because sham-controlled post-LIFU imaging was unavailable and one patient did not experience hallucinations prior to the sham sonication, these findings should be interpreted as preliminary and interpreted with caution. Furthermore, although expectancy/placebo effects cannot be fully ruled out, blinding integrity was high: no participant correctly identified the sham condition, and sensory experiences were reported as indistinguishable across sessions.

These limitations highlight the need for a larger, fully powered, sham-controlled, double-blind mechanistic clinical trial. Future studies should (i) incorporate real-time neurophysiological readouts such as fast fMRI to capture immediate circuit responses; (ii) refine individualized targeting using connectivity-based or symptom-linked functional markers; (iii) examine dose–response relationships and optimize LIFU temporal patterns; and (iv) evaluate durability of LIFU-induced symptom reduction across multiple sessions. Such designs will be essential to determine whether striatal LIFU can reliably modify hallucination-specific circuits and produce clinically meaningful, sustained reductions in hallucination severity.

### Conclusions

This first-in-human study demonstrates that neuronavigated LIFU can induce rapid reductions in auditory hallucination severity while safely and precisely modulating aberrant striatal–STC connectivity. These preliminary findings highlight the striatal–STC pathway as a compelling mechanistic target for noninvasive neuromodulation in psychosis and position LIFU as a uniquely transformative approach for precise, deep striatal circuit interrogation and modulation in severe psychosis-spectrum disorders. These results underscore the urgent need for larger sham-controlled trials to establish the causal role of striatal circuits in hallucination generation and to evaluate the therapeutic potential of LIFU for treatment-resistant schizophrenia.

## Disclosures

EDK, JLA, RM, and KRM hold equity in and are paid a salary by Attune Neurosciences Inc.

## Acknowledgments

We thank all the participants for completing our studies. This research is supported by NIMH R01 grants (R01MH122897 and R01MH122897-05S1) to Karuna Subramaniam.

## Supplemental Methods

### Individualized Transcranial Acoustic Simulations

Before stimulation, individualized transcranial acoustic simulations were performed for each SZ participant to estimate in situ pressure, intensity, and temperature distributions at striatal targets (NAc, CH). Simulations incorporated participant-specific skull geometry derived from pseudo-CT images generated from zero-TE (PETRA) MRI sequences (61). Simulation source pressures were calibrated for each 64-element array across a 3D dimensional steering space using a capsule hydrophone (ONDA HGL-400) calibrated at 500 kHz mounted in the ONDA AIMS III scan tank (Product # AST3-L-3). Directivity loss compensation was calculated using a Bessel function fit as specified by the manufacturer. Water was degassed and filtered continued with an Aquas-10 Water Conditioning system (Onda product # AQUAS-10-110V). These models accounted for inter-individual variability in skull thickness, density, and acoustic attenuation. All simulation outputs remained below the ITRUSST biophysical safety limits (MI < 1.9; ΔT < 2°C, see Table 2) (64–66). Additional 95^th^ percentile error estimates of brain mechanical index were included to account for worst-case acoustic simulation error when transmitting through bone.

### Neuronavigation and Targeting Procedure

Participants underwent individualized MRI-guided neuronavigation using the Attune transducer and navigation software. Each participant’s high-resolution T1-weighted anatomical MRI and pseudo-CT (PETRA-derived) images were imported to estimate skull geometry, local bone density, and acoustic propagation characteristics to optimize beam alignment. Striatal targets were defined as 5-mm radius spheres centered at canonical coordinates: left NAc: MNI −8, 9, −8, right NAc: MNI 8, 9, −8, left caudate head (CH): MNI −10, 18, 2, and right caudate head (CH): MNI 10, 18, 2. These coordinates were transformed to each participant’s native anatomical space, and the resulting target locations were used to guide transducer placement with millimeter precision.

Prior to sonication, hair was parted and conductive ultrasound gel was applied to ensure optimal acoustic coupling and eliminate air gaps. The 128-element 500 kHz transducer was positioned to align the beam path with the skull’s local surface normal, minimizing refraction and maximizing transmission to the deep striatal foci (depths: ∼70 mm for NAc; ∼65 mm for CH). Attune software computed patient-specific beam trajectories and overlaid focal locations directly onto the subject’s MRI to verify accurate targeting. Transducer coordinates were recorded at the beginning and end of each sonication session to confirm targeting accuracy. Personalized transcranial acoustic simulations were used to confirm that focal energy deposition was confined to the intended striatal target and that predicted pressure and thermal indices remained within FDA and ITRUSST safety limits. All participants showed stable beam alignment throughout each sonication session, with no evidence of drift or off-target energy deposition. Beam trajectories consistently converged at the striatal targets (confirmed by modeling and MRI overlays).

Sonication parameters were: 20 Hz pulse repetition frequency, 6% duty cycle (3 ms pulses, 50 ms interval), applied in 40 s on / 20 s off cycles for 30 minutes per target site in each hemisphere (60 minutes total). This temporal pattern was selected to: minimize cumulative heating, maintain stable focal intensity, and adhere to ITRUSST thermal safety thresholds. All participants tolerated stimulation well; no participant reported adverse events beyond mild, transient sensations (e.g., faint auditory clicks or brief scalp tightness associated with transducer positioning).

### Sham Control Condition (Unfocused Sonication)

The sham condition used unfocused sonication (UF) designed to preserve peripheral sensory experiences while eliminating focal brain energy deposition. Given that the striatal targets lie approximately 65 mm from the transducer face, near field pressure distributions (e.g., around the temples) are relatively uniform. For the UF control, all procedures matched active LIFU, including transducer placement, but random time delays were applied across the 128-element array to eliminate constructive interference and prevent focal brain engagement. This configuration produces a diffuse planar pressure field that mimics the spatial pressure profile of the near field in the active condition. As a result, it preserves peripheral sensory experiences—such as subtle bone-conducted sounds (“clicks”) or scalp sensations. This design ensures that the UF condition replicated the auditory and somatosensory experience of active LIFU without engaging brain targets. All other procedures, including transducer placement, were identical to the active sessions. Ultrasound beam trajectories verified the absence of any focal brain hotspot during UF stimulation.

### Resting-State fMRI Preprocessing and Seed Definitions

Resting-State fMRI (rs-fMRI) preprocessing was performed using SPM12 and CONN v22. Steps included: slice-timing correction, realignment and motion correction, co-registration to native T1, segmentation and normalization to MNI space, spatial smoothing (6 mm FWHM), and denoising using aCompCor and motion regressors. Striatal regions of interest were defined as 5-mm radius spheres centered at: NAc: MNI −8, 9, −8 and CH: MNI −10, 18, 2. Seed-to-voxel connectivity was calculated using Fisher Z-transformed correlations between each seed and the rest of the brain, with focus on STC regions. The magnitude of Fisher Z-transformed correlations was computed between striatal seeds (NAc, CH) and STC, given prior evidence of striatal–temporal hyperconnectivity underlying auditory hallucinations (5, 6, 14, 30).

### Safety Monitoring

Safety monitoring included MI tracking and skull and brain temperature recordings during simulations and each sonication session. Real-time monitoring included: participant-reported sensations, and immediate session termination if moderate discomfort occurred. No moderate or severe adverse events occurred.

## Data Availability

All simulation parameter sets, beam models, targeting coordinates, and preprocessing scripts will be made available upon request or via public repository upon acceptance.

---

**Supplemental Figure 1.**
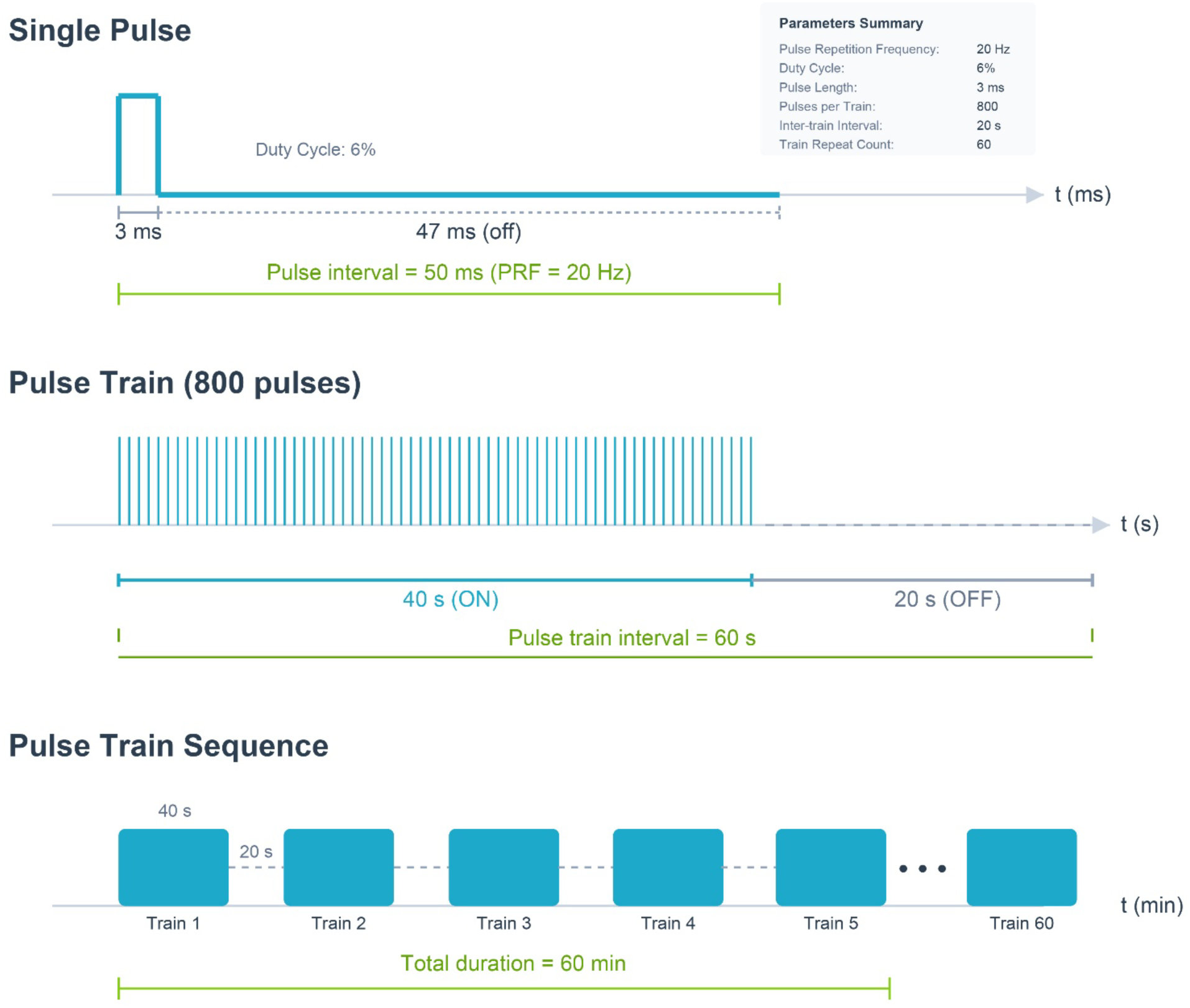
Ultrasound stimulus parameter diagram

## Notes

### Clinical Trial

NCT04807530

### Author Declarations

This study was conducted under an NIMH-funded R01 (R01MH122897) to Karuna Subramaniam and designated non-significant risk (NSR) by the UCSF Institutional Review Board (IRB #24-42726). SZ were recruited via ClinicalTrials.gov (NCT04807530) and provided informed consent for this protocol approved by Attune Neurosciences and the IRB at UCSF prior to study enrollment.

